# Patient Perspectives and Satisfaction: Educational Needs and Communication Barriers in Dermatology Clinics in Saudi Arabia – A Cross-Sectional Survey

**DOI:** 10.64898/2026.03.11.26348118

**Authors:** Fawwaz Freih Alshammarie, Aala Hazza Alhobera, Maryam Abdullah Alshammari

## Abstract

**Purpose:** Understanding patient perspectives is essential to improving quality and satisfaction of care in dermatology clinics. In Saudi Arabia, limited national data exist on patients’ educational needs and communication barriers. This study aimed to assess patient satisfaction, identify educational gaps, and explore communication challenges in dermatology clinics across Saudi Arabia.

**Patients and methods:** A national cross-sectional survey was conducted among 976 dermatology patients. A structured questionnaire evaluated demographics, perceived knowledge, satisfaction with information provided, communication barriers, and preferred educational methods. Descriptive statistics and chi-square tests were used for analysis.

**Results:** Among participants who had attended dermatology clinics (n = 795), 61.6% reported frequent or occasional confusion about their condition, and only 45.4% demonstrated high self-reported knowledge. Overall satisfaction was moderate, with 58.3% satisfied or very satisfied, while 9.9% reported dissatisfaction. The most reported communication barriers were limited consultation time (25.2%) and patient anxiety about asking questions (15.3%). Patients felt least informed about treatment options (22.6%), diagnosis (20.3%), and potential side effects (19.3%). Most participants (70.6%) preferred language communication to be in Arabic, and 78% favored the physical method of face-to-face education consultation. Patient knowledge, barriers and preferences significantly differed with age, gender, and condition complexity (p < 0.05).

**Conclusion:** Dermatology patients in Saudi Arabia report moderate satisfaction with substantial educational needs and communication barriers. Addressing consultation time constraints, fostering supportive communication environments, and providing patient-centered, language-appropriate education; particularly through direct face-to-face interactions will aid to enhance understanding, satisfaction, and engagement in for an overall better provider-patient dermatologic care.

## Introduction

Patient education is an essential component of dermatologic care and plays a critical role in improving treatment adherence, patient satisfaction, and clinical outcomes. Effective education of patient by health care provider enables them to understand their conditions, apply treatments correctly, and engage in self-management behaviors that support long-term disease control. Evidence from chronic dermatologic conditions, including atopic dermatitis, demonstrates that structured educational interventions improve patient comprehension, engagement, and adherence to medical recommendations [1]. Despite these benefits, many dermatology patients report gaps in the clarity, accessibility, or adequacy of the information they receive in clinical settings [2].

Variability in the delivery of patient education is influenced by consultation time constraints, diverse patient learning preferences, and differences in health literacy. Studies highlight that many standard patient education materials are not fully adapted to patient needs, and personalization, such as tailoring communication to language, preferred format, and cultural context, all of which enhance satisfaction and understanding [3,4,5]. Patients’ preferred methods for receiving information often vary; while digital resources are increasingly available, face-to-face interactions remain the most valued approach, providing reassurance, clarity, and opportunities for questions [5,6,7].

Patient comprehension is also shaped by individual and condition-related factors, including age, gender, prior confusion, and the complexity of the skin condition. For example, confusion or anxiety can negatively affect understanding, whereas patients with less complex conditions often demonstrate higher knowledge [1,5,6]. Additionally, patients frequently seek supplemental information outside clinical encounters, commonly via the internet, which underscores the importance of providing reliable, clinic-based educational resources to prevent misinformation [7]. In the Kingdom of Saudi Arabia, only limited research has explored dermatology patient education from the patients’ perspective. Understanding current educational practices, perceived adequacy of information, knowledge gaps, and patient preferences is essential to guide the development of evidence-based, patient-centered strategies. The cross-sectional survey presented in the current study aims to provide an overview of patient education in dermatology clinics in Saudi Arabia, examining formats, content, perceived adequacy, and preferred educational methods. By identifying unmet needs, the study seeks to identify effective strategies that enhance understanding, satisfaction, and patient involvement in dermatologic care.

## Material and methods

### Study Design and Participants

A comprehensive national survey across Saudi Arabia was conducted for the study, targeting 976 participants all within the age of 18 years and above from both male and female gender through the method of convenient stratified sampling. Majority of participants were aged 20-30 years old (445) (45.6%), with a higher proportion of female gender (786) (80.5%). Most of which were of a higher educational background; bachelor’s degree (582) (59.6%). The survey covered all regions of Saudi Arabia, with a larger percentage of respondents from Northen region (495) (50.7%). Stratifying data by age and gender to improve the inclusivity, thereby ensuring results to be of a diverse epidemiological population.

### Survey Method and Data

The survey utilized a structured anonymous questionnaire designed to collect both quantitative and qualitative data. It was developed based on a comprehensive review of the literature and consultations with senior dermatology physicians to ensure relevant aspects of patient education were adequately addressed. It collected demographic information, including age, gender, educational level, and region of residence. Relevant clinical history was also obtained, such as prior dermatology clinic visit, and skin condition participants were following for. More importantly, the questionnaire focused on exploring elements related to provider-patient communication and education, including identifying patients’ knowledge gaps and barriers, preferred methods and factors for a better patient understanding. The full questionnaire is provided in the supplementary materials.

### Study Setting, Period and Data Collection

The study was conducted by the College of Medicine, University of Hail, Saudi Arabia. Data collection was carried out over a two-month period, from 1^st^ of January till the 28^th^ of February 2025. Participants were recruited online via social media platforms and physically through public locations, such as public parks, malls and university campuses. Survey data last accessed by authors for research purposes on the 15^th^ of May 2025. Authors with access to survey data pose minimal risk of identifying individual participants as implemented survey was strictly anonymous ensuring a high level of participant anonymity.

### Statistical Analysis

Data were analyzed using Statistical Package for Social Studies for Windows, Version 26 (SPSS; Released 2019; IBM Corp., Armonk, NY, USA). Descriptive statistics, including frequencies and percentages, were employed to summarize the demographic and clinical characteristics of the study population. Categorical variables were analyzed using the Chi-square test to evaluate associations and identify statistically significant relationships between variables. When the expected cell count was <5 in 20% or more of the cells, Fisher’s exact test was applied to ensure the accuracy of the results. Statistical significance was determined at a p-value threshold of <0.05.

### Informed Consent

A waived informed consent in written format was obtained from all participants prior to completing an anonymous questionnaire, confirmed by either answering agree to participate or do not agree to participate in the study.

### Ethical approval

The study complied with ethical standards outlined in the Declaration of Helsinki. The study has been reviewed and approved by the Research Ethics Committee (REC) at University of Hail, College of Medicine. Dated: 02/12/2024 with the reference number of H-2024-507.

## Results

### Demographic and clinical characteristics

Table 1 presents the demographic and clinical characteristics of the study participants. A total of 976 individuals were included, with females representing the majority at 786 (80.5%) and males accounting for 190 (19.5%). The largest age group was 20–30 years, comprising 445 participants (45.6%), followed by 151 participants between 40–50 years (15.5%), 137 participants less than 20 years (14.0%), 134 participants between 30–40 years (13.7%), 85 participants between 50–60 years (8.7%), and those aged 60 years and above representing about 24 (2.5%). Regarding education, 582 participants held a bachelor’s degree (59.6%), while high school education was reported by 275 (28.2%), 56 participants held a master’s degree (5.7%), 44 participants reported secondary education (4.5%), and primary education was reported by 19 (1.9%). Geographically, participants were predominantly from the northern region 495 (50.7%), followed by the western region 185 (19.0%), middle region 178 (18.2%), eastern region 96 (9.8%), and southern region 22 (2.3%). Most participants had previously visited a dermatology clinic 795 (81.5%), while 181 (18.5%) had not and were therefore excluded from the study. Among those who had visited a clinic, with 318 (40.0%) reporting only one visit, 232 (29.2%) visiting once a year, 157 (19.7%) visiting 2–3 times a year, and 88 (11.1%) visiting more than three times annually. Regarding skin conditions, acne was the most common 427 (53.2%), followed by eczema 184 (22.9%), skin infections 63 (7.9%), rosacea 47 (5.9%), warts 33 (4.1%), psoriasis 18 (2.2%), and other conditions 30 (3.7%), with multiple responses allowed.

**Table 1.**
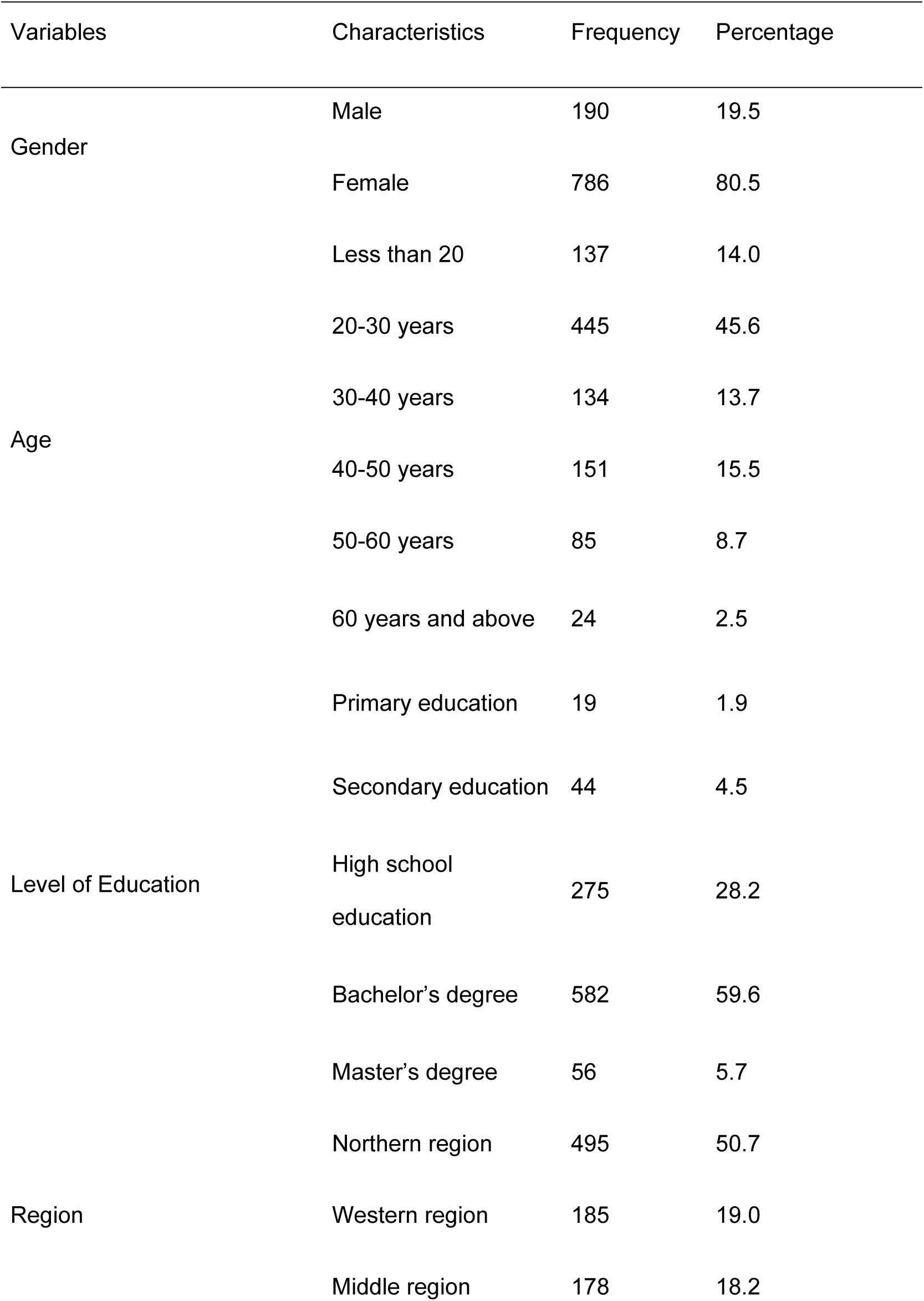
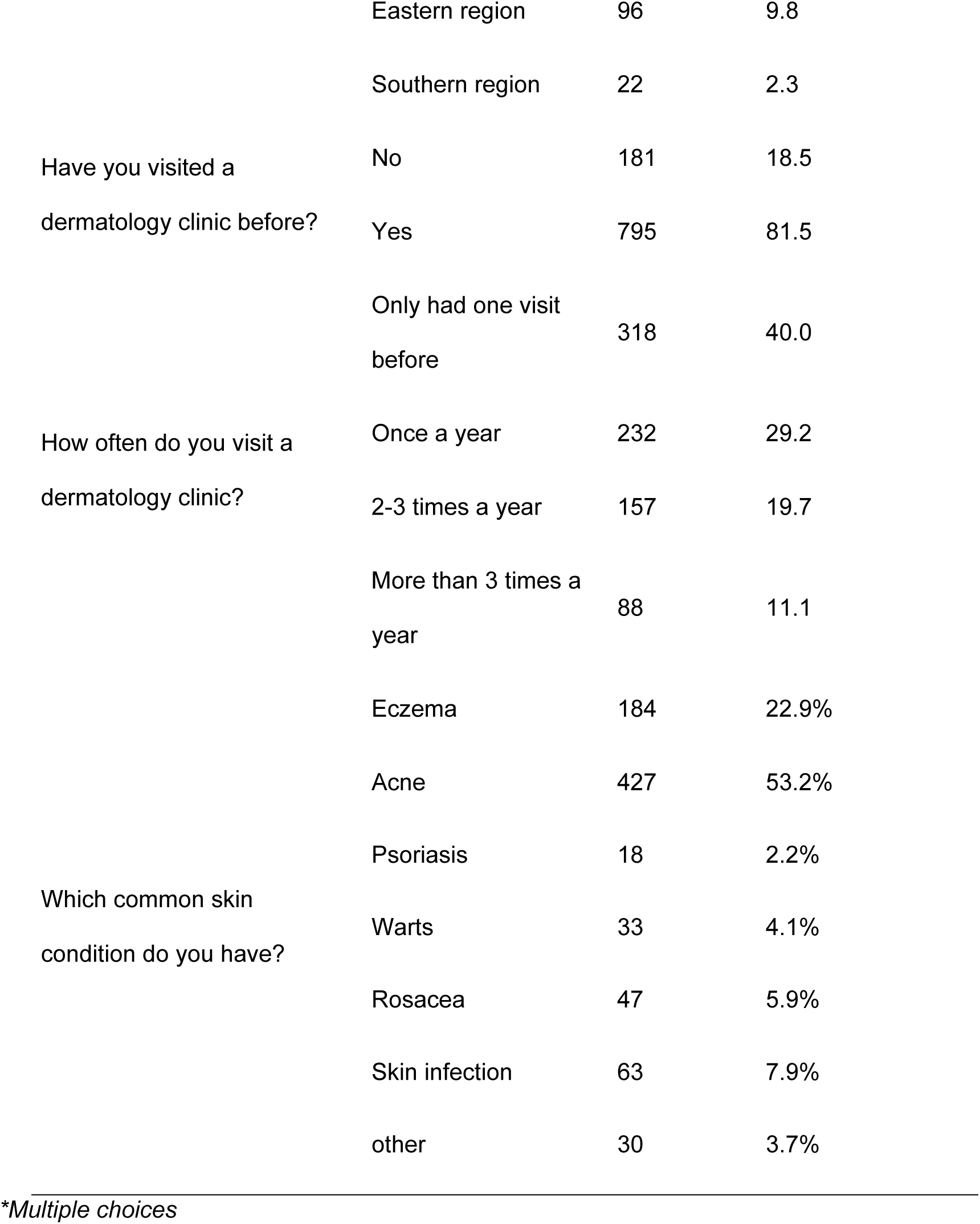
Demographic and clinical characteristics.

### Patients’ knowledge gaps regarding their skin conditions

Table 2 depicts an overview of patients’ knowledge gaps regarding their skin conditions. Among participants ((N = 795), 133 (16.7%) of respondents reported often being confused or misunderstanding information about their skin condition, 357 (44.9%) experienced some confusion, 170 (21.4%) rarely felt confused, and 135 (17.0%) never felt confused. When asked how much general knowledge they felt they had about their condition at the current time, 161 (20.3%) reported low knowledge, 273 (34.3%) reported moderate knowledge, and 361 (45.4%) reported high knowledge. Regarding the complexity of the dermatology skin condition role in understanding their condition, 443 (55.7%) believed their understanding was not affected, 122 (15.3%) thought complexity affected their understanding, and 230 (28.9%) were neutral regarding this aspect. When asked if they were satisfied with the information provided by their dermatologist, 152 (19.1%) were very satisfied, 312 (39.2%) were satisfied, 252 (31.7%) were neutral, 56 (7.0%) were not satisfied, and 23 (2.9%) were not satisfied at all. Patients shared many barriers that impeded their understanding at their clinic visit including: not having enough time with their provider at 313 (25.2%) of patients, feeling anxious or scared to ask their questions at 190 (15.3%) of patients, thinking the provider was too busy at 121 (9.8%) of patients, caregivers at the visit could not communicate in the same language as doctor at 59 (4.8%) of patients, having medical terminology that was impossible to understand at 66 (5.3%), other barriers at 52 (4.2%) of patients, and finally, 439 (35.4%) of patients did not have barriers. Regarding medical aspects about which patients felt least informed, 248 (20.3%) indicated diagnosis, 80 (6.6%) causes and risk factors, 276 (22.6%) treatment options, 159 (13.0%) medication usage, 200 (16.4%) prevention and lifestyle measures, 235 (19.3%) potential side effects or complications, and only 22 (1.8%) felt fully informed.3.2. Figures, Tables and Schemes.

**Table 2.**
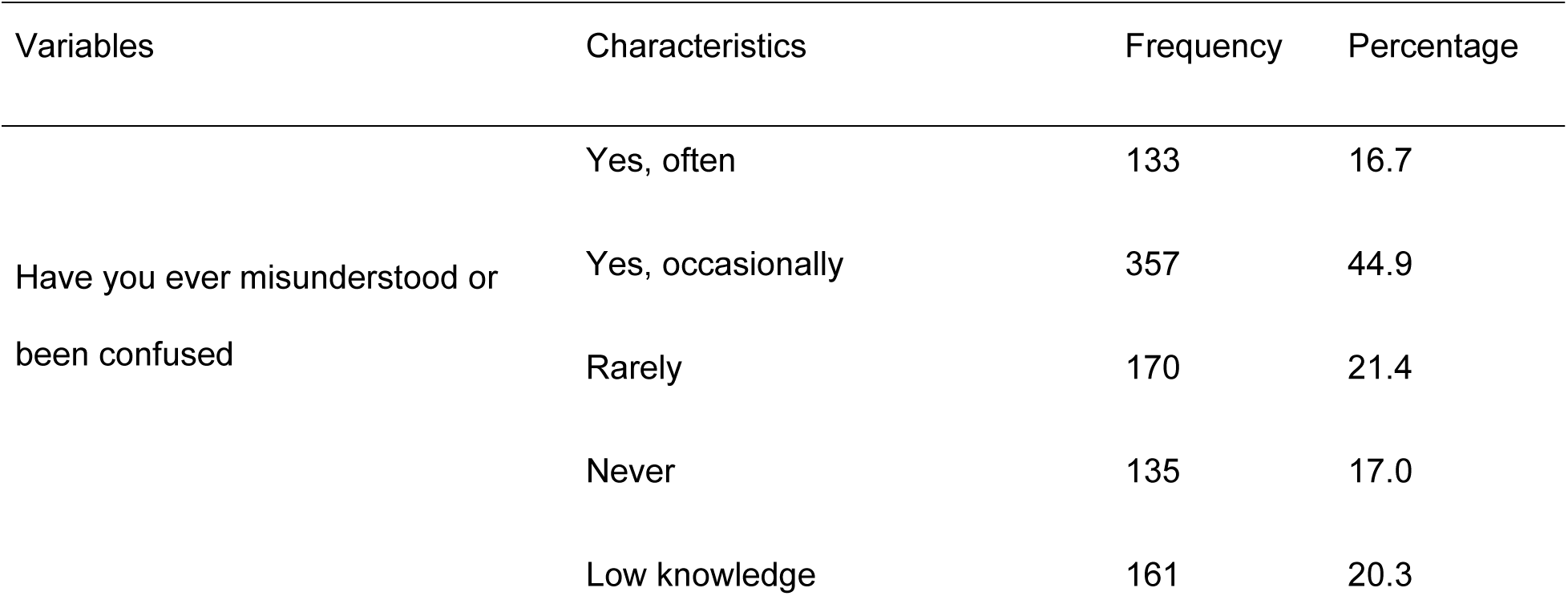
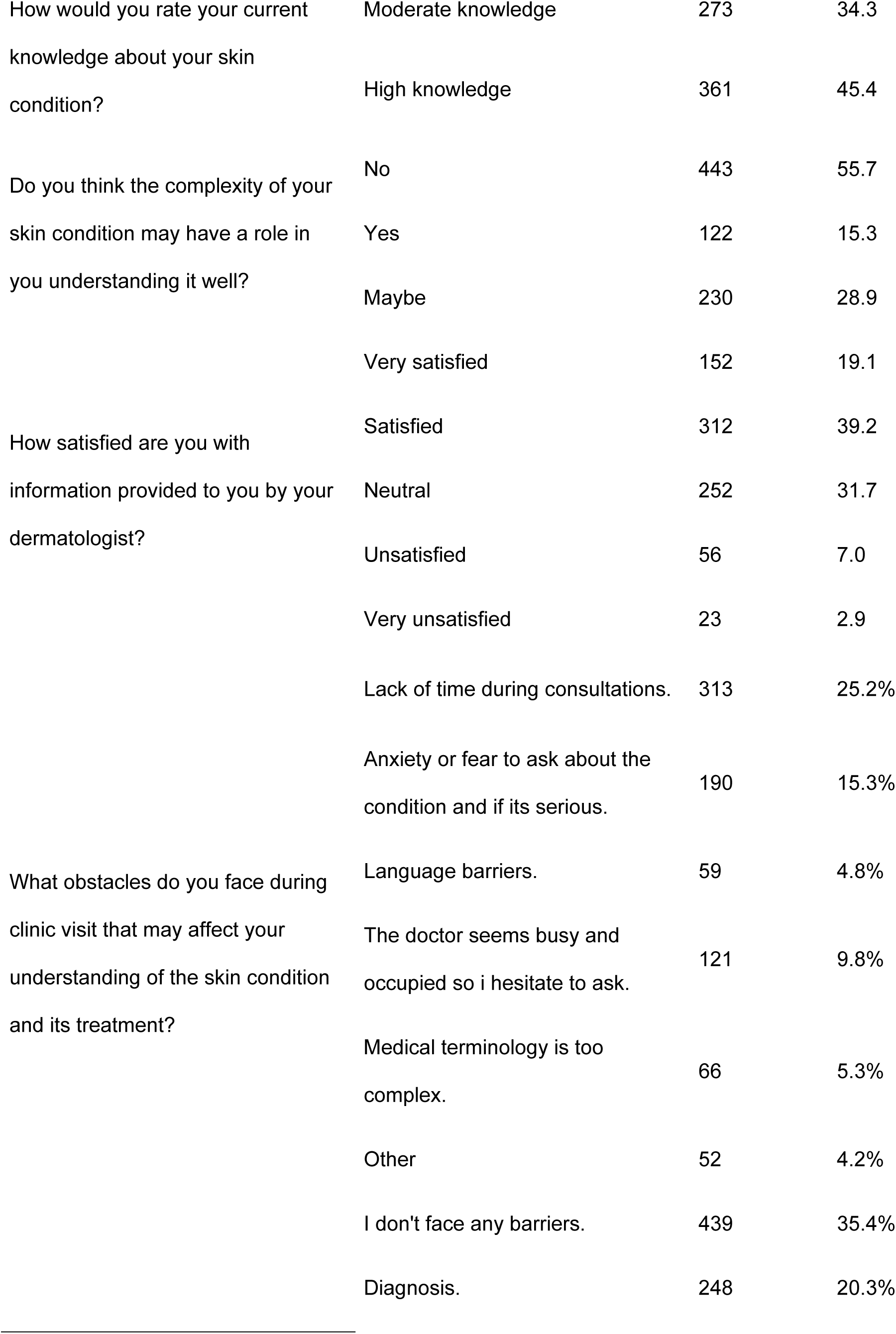
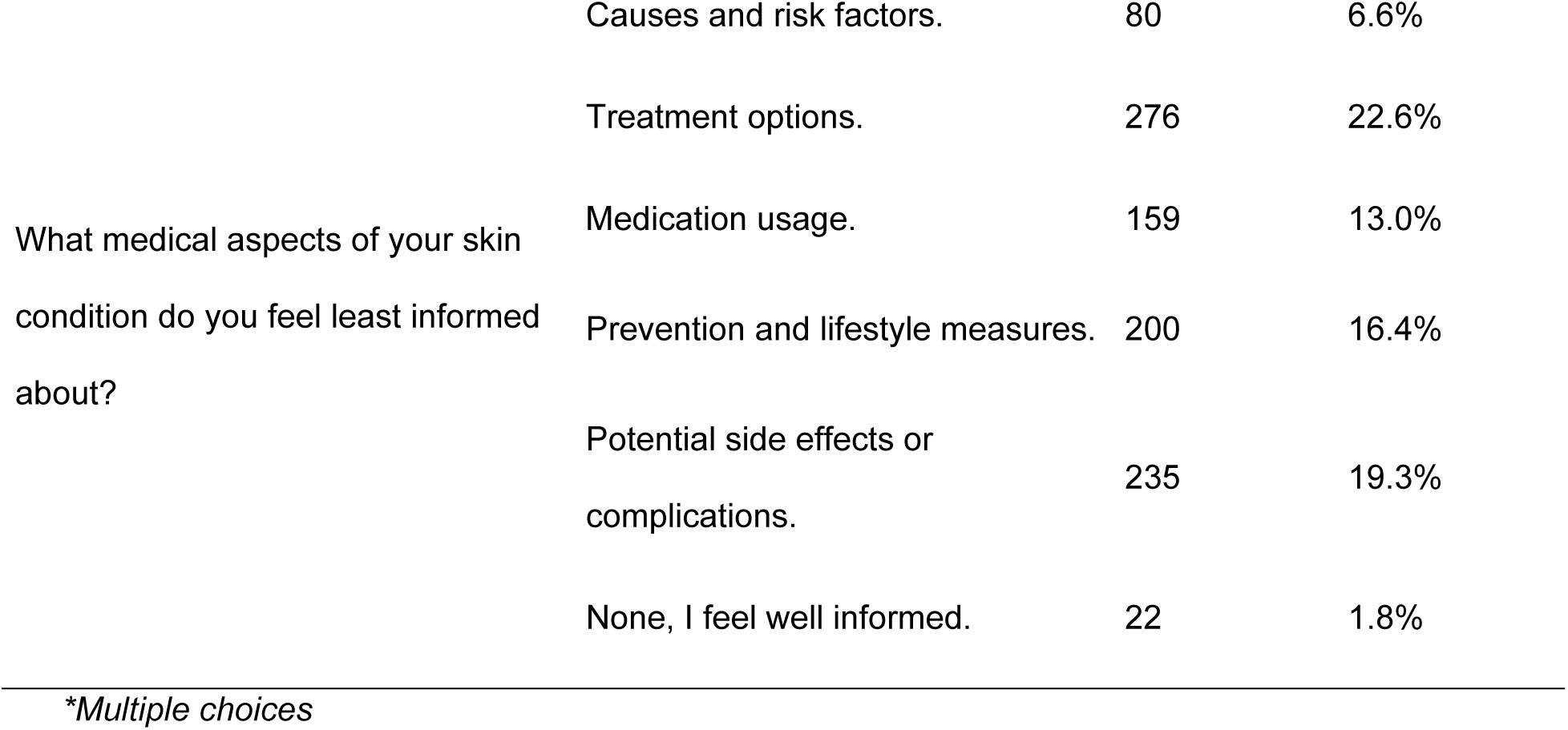
Patients’ knowledge gaps regarding their skin conditions.

### Patients’ preferences for receiving information

Table 3 presents an overview of patients ’preferences for receiving information about their skin conditions. The majority of participants (70.6%) preferred to communicate with their provider in Arabic as they understood it better, while 5.7% preferred English since most information online and names of skin conditions are in English, and 23.8% expressed no preference for either language. When asked if they had ever sought information outside of their clinic visits because they felt confused or that the information provided was not enough, most respondents (72.2%) reported doing so, whereas 27.8% had not. Among those who sought additional information, online sources such as social media and websites were the most common (57.1%), followed by advice from family and friends (25.6%), support groups either in person or online (10.8%), and books or pamphlets (6.4%).

Figure 1 presents participants’ preferred methods of receiving information about their skin condition, face-to-face consultations were overwhelmingly favored as the first choice by 620 participants (78%), with smaller proportions selecting it as their second 84 (10.6%), third 45 (5.7%), or fourth choice 24 (3%), and only 11 participants (1.4%) ranking it fifth or sixth. Digital written sources, such as websites and Twitter, were chosen as the first preference by 202 participants (25.4%) and were most commonly selected as the second choice 245 (30.8%). Video platforms like YouTube and TikTok were preferred as the first choice by 220 respondents (27.7%), followed closely by group sessions, chosen as the first choice by 141 participants (17.7%) but more frequently ranked in the fourth to sixth positions. Printed materials were less frequently selected as the first choice 99 (12.5%) but gained higher preference in the second and third positions (181; 22.8% and 169; 21.3%, respectively). Finally, other methods were selected as the first choice by 192 participants (24.2%), though the majority favored them as a sixth choice 268 (33.7%). Overall, face-to-face consultations clearly dominated participants’ preferences, while digital, video, and group-based methods showed more varied rankings across the sample.

**Figure 1:**
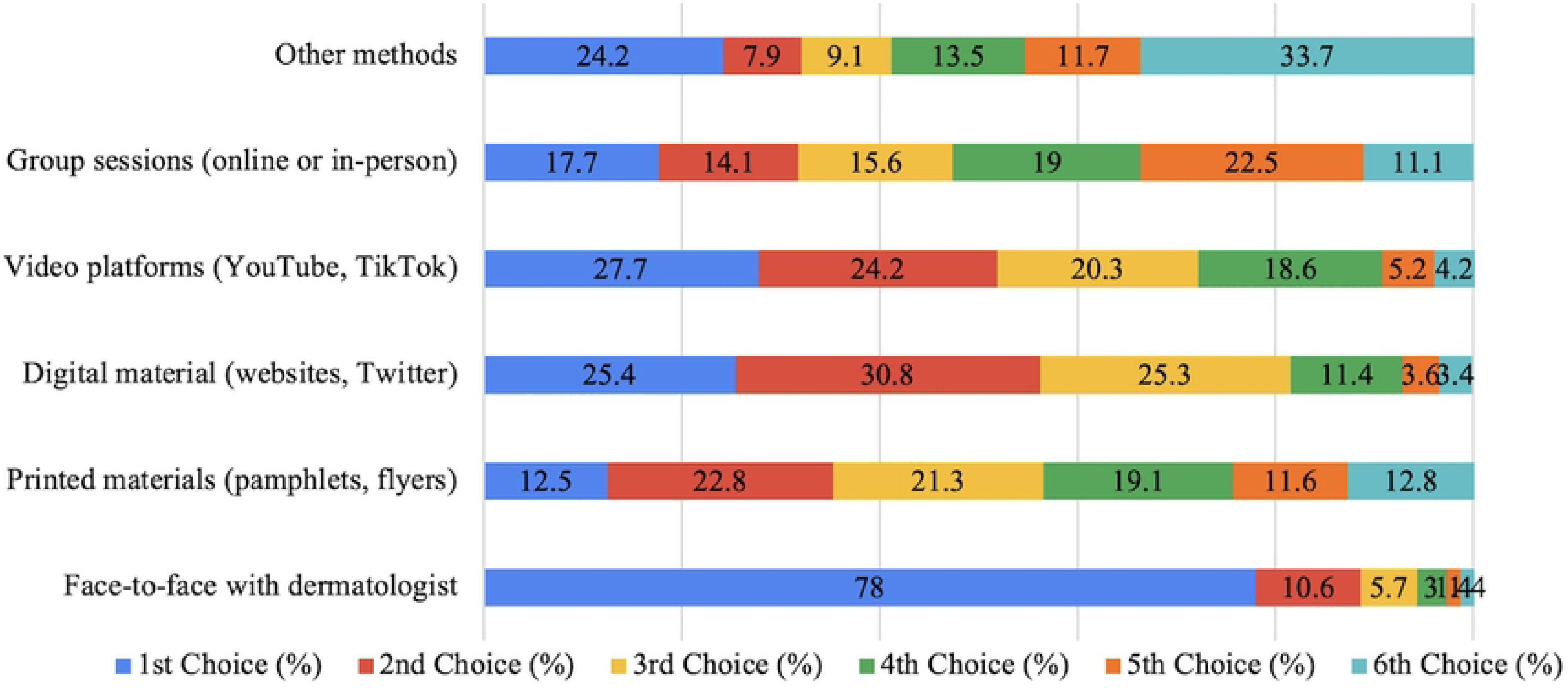
Preferred Methods of Receiving Information (N = 795)

**Table 3.**
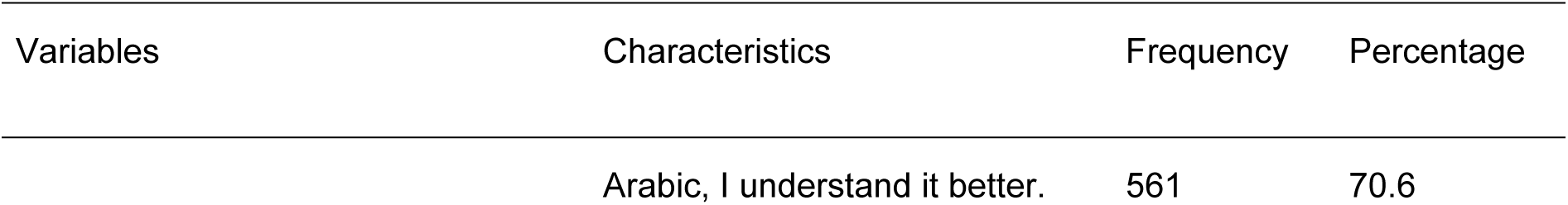
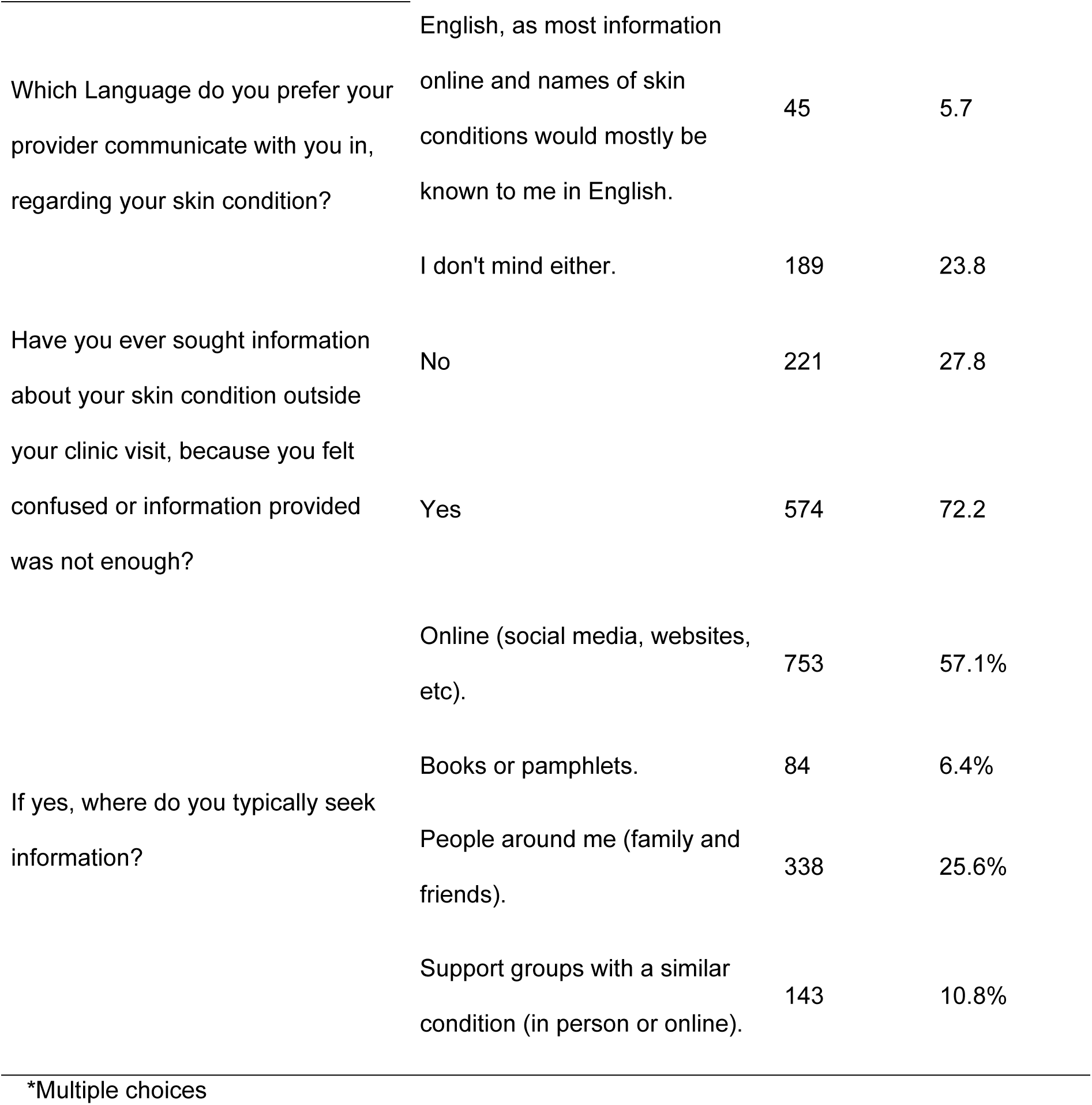
Patients ’preferences for receiving information about their skin conditions.

Table 4 presents an overview of patients **’**perceptions regarding factors that improve understanding of their skin condition and their engagement with educational resources. Among participants (N = 795), step-by-step guides for managing their condition were considered the most helpful by 796 (35.0%), followed by closure summaries provided by the doctor at the end of the visit by 264 (11.6%), visual aids in the clinic such as 3D skin models or diagrams by 231 (10.2%), more time spent with the dermatologist during each visit by 224 (9.9%), patients **’** success stories or case study examples (219, 9.6%), contact information for additional support like nurse hotlines or support groups by 203 (8.9%), research updates and new treatment options by 182 (8.0%), and availability of a patient educator to provide further explanations and support by 153 (6.7%). Regarding whether participants felt their dermatologist took enough time to answer questions and explain their condition, 257 (32.3%) reported **“**yes, always,” 297 (37.4%) stated**“** yes, most times,” and 241 (30.3%) reported**“** sometimes.” When asked about how often they would like updates about their skin condition, 406 (51.1%) preferred updates at every visit, 93 (11.7%) once a month, 86 (10.8%) every three months, 170 (21.4%) only when there were significant changes, and 40 (5.0%) did not care to be informed. Regarding participation in educational workshops or webinars hosted by the clinic to help other patients and improve awareness, 342 (43.0%) expressed interest, 275 (34.6%) were unsure, and 178 (22.4%) were not interested. Finally, when asked if they would be willing to provide feedback on educational materials to improve them, 501 (63.0%) answered yes, while 294 (37.0%) declined.

**Table 4.**
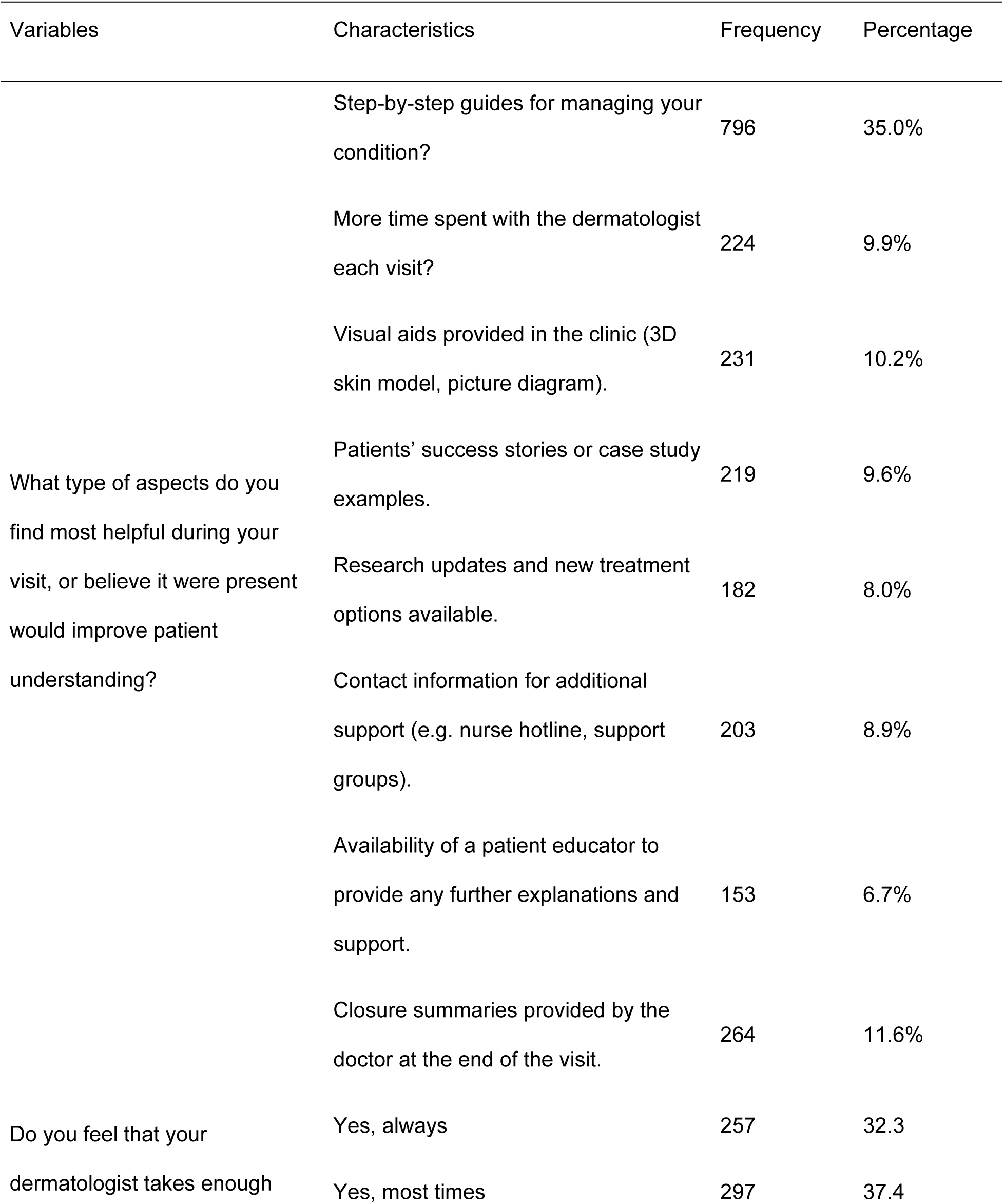
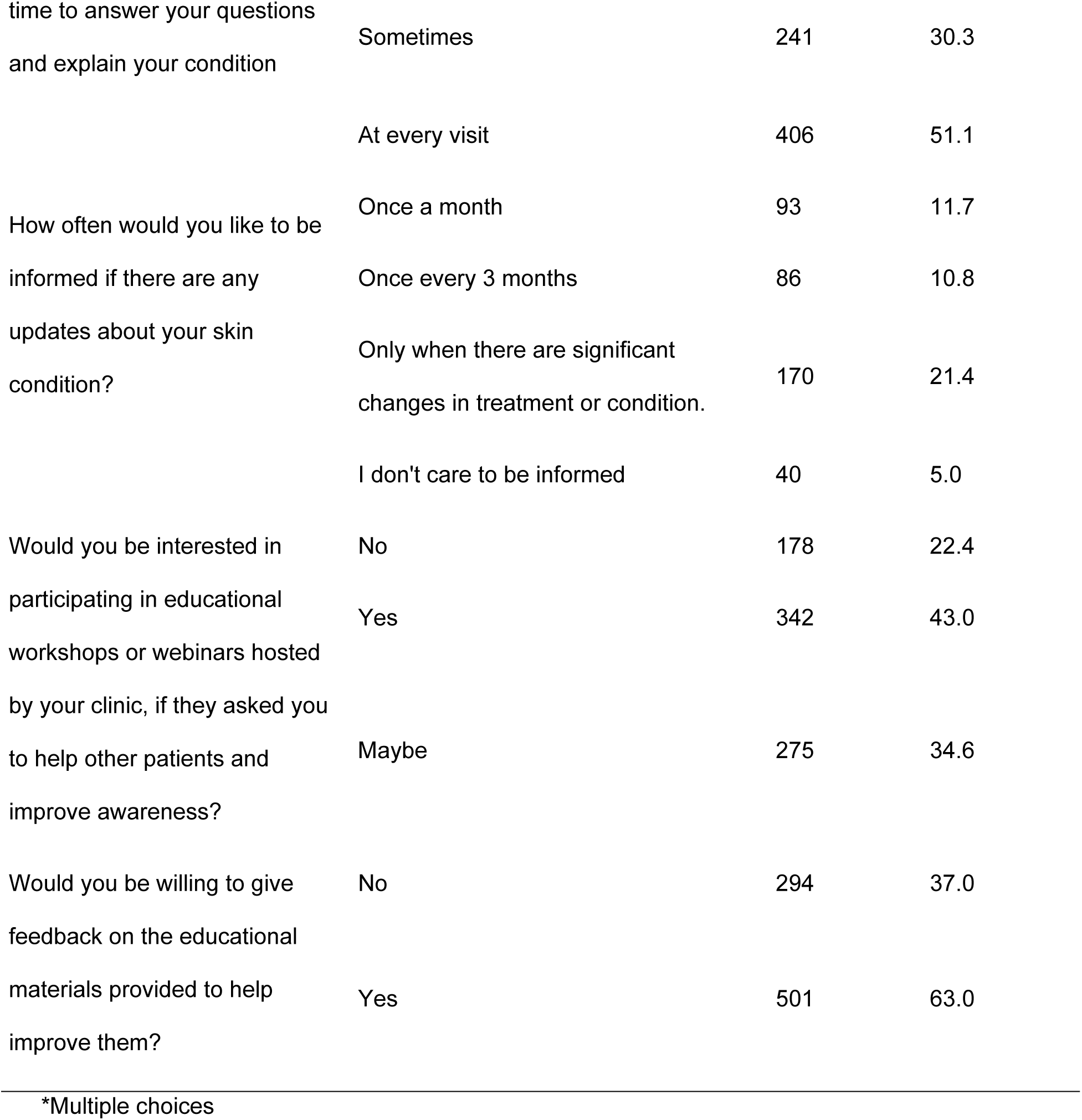
Patients ’perceptions regarding factors that improve understanding of their skin condition.

The results presented in Table 5 illustrate the associations between participants’ characteristics and their knowledge about their skin condition. Male participants were slightly more likely to have high knowledge 72 (49.0%) compared with females 289 (44.6%) (χ² = 6.446, p = 0.040). Age also showed a significant association, with participants aged 40–50 years demonstrating the highest proportion of high knowledge 75 (57.3%), while those under 20 years had the lowest 42 (43.3%) (χ² = 19.959, p = 0.030). Although education level appeared related to knowledge, the association was not statistically significant; participants with a master’s degree had the highest proportion of high knowledge 27 (56.3%) compared with those with primary education 8 (44.4%) (χ² = 7.033, p = 0.533). Regarding misunderstandings about their condition, those who reported rarely and never being confused had markedly greater in high knowledge levels 102 (60.0%) and 97 (71.9%), respectively, compared with participants who often experienced confusion 40 (30.1%) (χ² = 95.993, p < 0.001). The complexity of the skin condition was strongly associated with knowledge: participants without complex conditions were more likely to have high knowledge 238 (53.7%) than those with complex conditions 29 (23.8%) (χ² = 70.793, p < 0.001). These findings emphasize that age, gender, prior confusion, and condition complexity significantly influence patients’ understanding of their skin health.

**Table 5.**
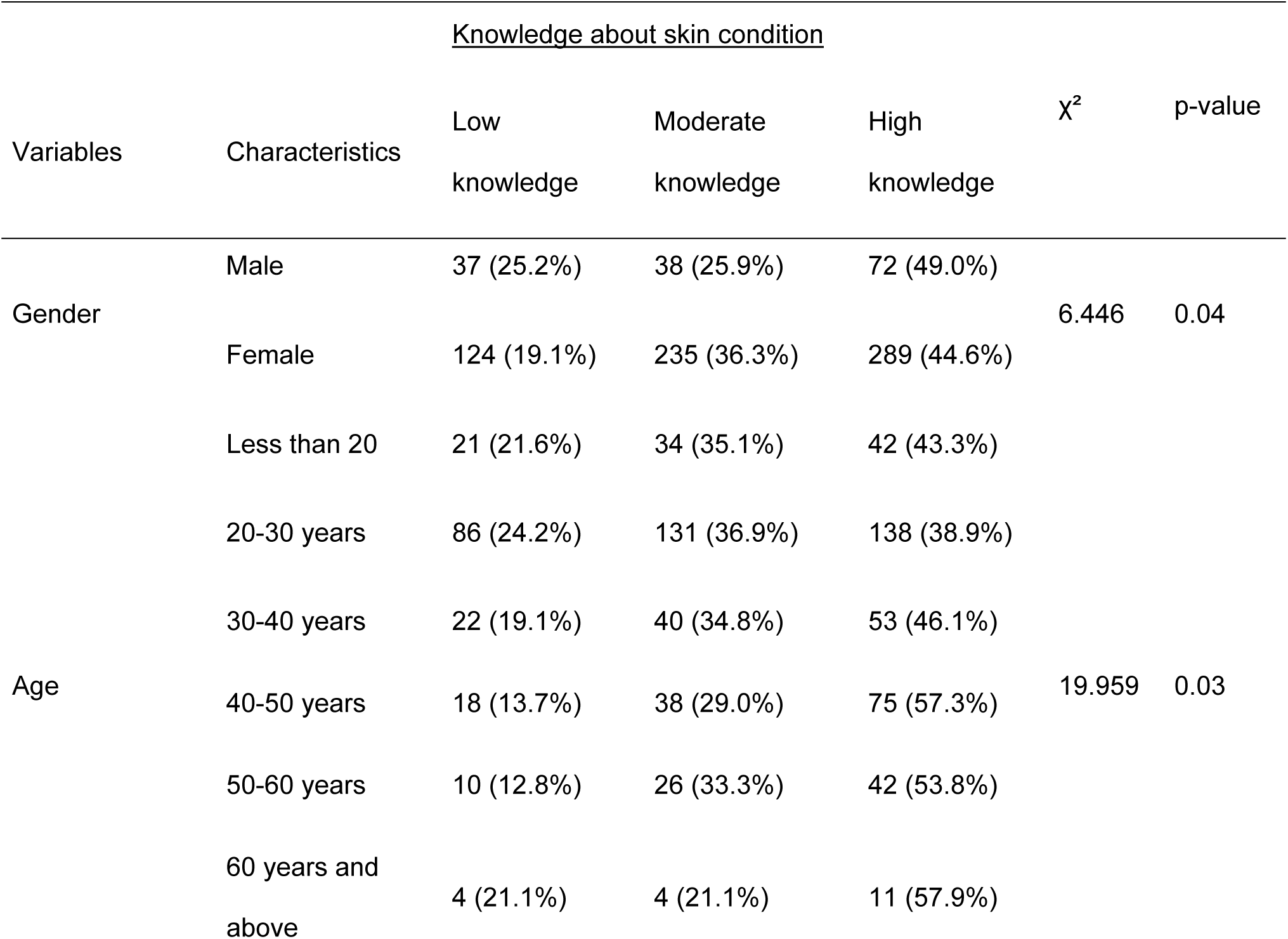
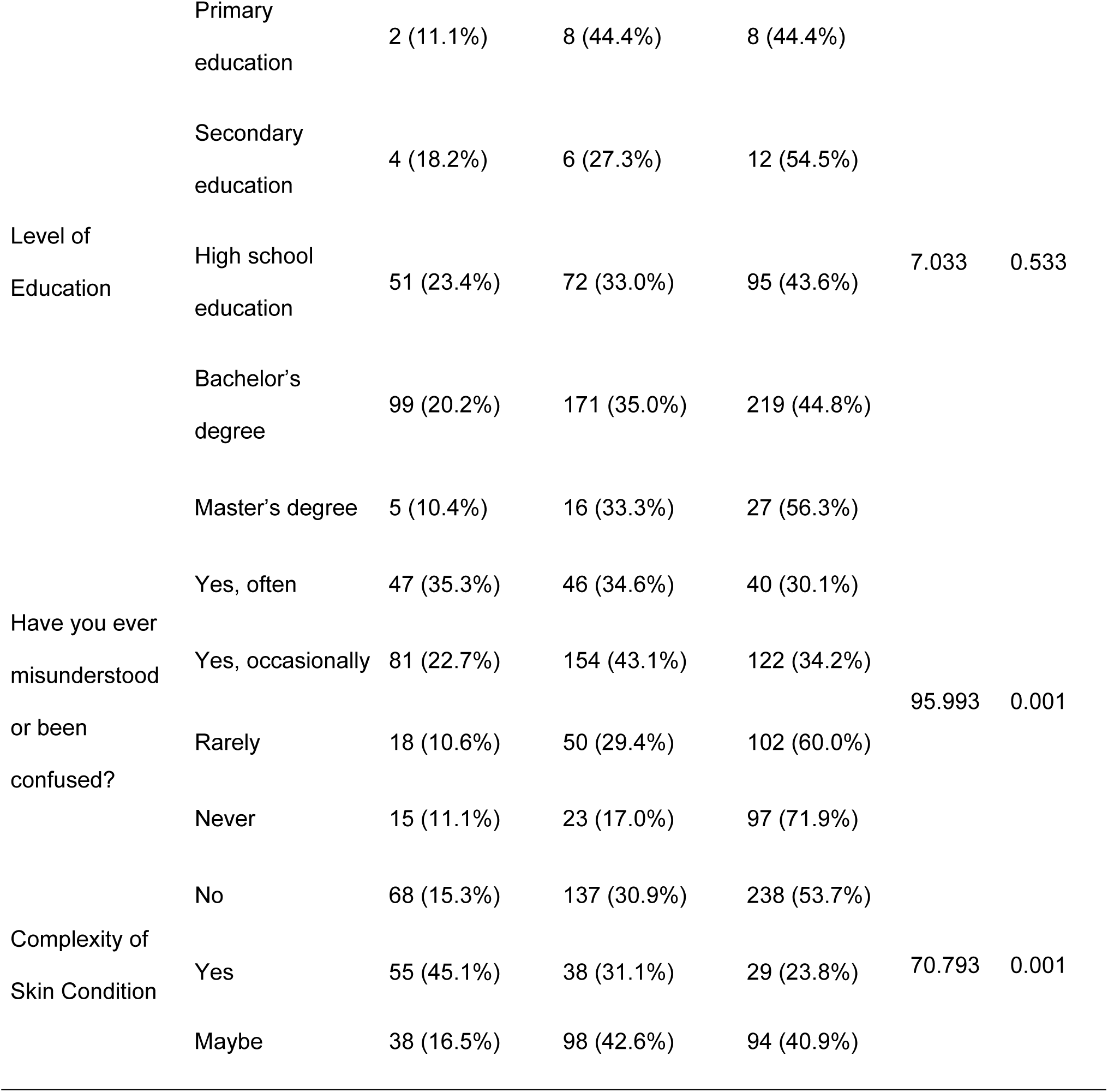
Associations between participants’ characteristics and their knowledge about their skin condition.

Participants were asked what improvements they would suggest to enhance patient education in dermatology clinics. Based on their responses, five key suggestions were identified to improve patient education in dermatology clinics. The first suggestion is to provide ample time for each patient with in-depth explanation of the patient’s condition, treatment options, potential side effects, and preventative care. The second suggestion is to improve communication by using straightforward, unambiguous language, visuals, and interactive resources so patients better understand their diagnosis and clinical care plan. The third suggestion is to provide patients with personalized educational materials at numerous levels, and some follow-up support such as digital resources, virtual consultations, and reminders during support group meetings. The fourth suggestion is to ensure physicians show empathy, active listening, and psychological support to patients, and create a supportive environment for patients to feel they have some agency in asking questions and sharing concerns. The fifth suggestion is to improve accessibility and cost-effectiveness of services for dermatology patients by providing additional appointments, making that care more reasonably available, or even advising on feasible and economical treatments.

## Discussion

This study evaluates the importance of patient education in medical settings in the Kingdom of Saudi Arabia by looking from the patients’ perspectives, asking for suggested improvements, and considering the feasibility of measuring education effectiveness. There were evident knowledge gaps between patients and their skin condition. Almost 490 (62%) of participants indicated some confusion about their condition, and less than half 361 (45.4%) identified having high knowledge. Whereas the majority of patients were of the opinion their condition did not complicate understanding their diagnosis, the level of satisfaction with the information received was moderate, with 252 (31.7%) neutral, and only 152 (19.1%) of patients identifying being very satisfied with the education they received. A frequent barrier to understanding the medical education provided was limited time with a medical provider (313 (25.2%)) and anxiety about asking questions (190 (15.3%)). Patients identified feeling least informed about their management options (276 (22.6%)), the diagnosis itself (248 (20.3%)), and possible treatment side-effects (235 (19.3%)). These represent important opportunities where medical education could be improved. Ultimately, study findings suggest that improved patient education strategies and communication with patients will address important knowledge gaps and support improved patient understanding, satisfaction, and involvement in their treatment and management of their skin condition. The findings of this study corroborate previous literature that highlights the significance of patient education in dermatology. A recent study [8] reported that barriers to understanding, poor recall, and the physician’s delivery are some of the main barriers to effective patient education, similar to the confusion and moderate satisfaction reported in our study. Another study [9] found that, even with regular follow-ups, there were significant knowledge gaps in disease management and treatment misconceptions, reinforcing the need for targeted education. Clearly, the topics in which patients felt least informed included treatment options like the use of micrographic surgery (MoHs), diagnosis, and possible side effects, which are typical areas of deficiencies reported in dermatology populations and requires systematic educational strategies to enhance knowledge, patient satisfaction, and active engagement in self-care. The study results identified patient preference for communication in Arabic (562, 70.6%) and noted that it was common for patients to seek additional information beyond their visits (574, 72.2%) - primarily the internet as a source (454, 57.1%). Additionally, close to 80% (620, 78.0%) of participants indicated that face-to-face encounters was the most preferred way to receive information, while ’digitally’, ’video’, or ’group’ orientated methods tended to be more variable in responses. Overall, the findings suggest that in-person interaction continues to be the most valued approach to patient education, despite the existence of alternative information sources. This preference for direct, in-person interactions echoes study [10], which reported that patients valued access to dermatology clinic notes and felt it enhanced understanding and involvement in care. Additionally, a study [11] highlighted that while social media can provide curated educational content, it may also spread misinformation, which could explain patients **’**reliance on direct consultations despite the availability of online sources. The findings suggest that patient education strategies must balance the convenience of digital resources with the personalized reassurance of face-to-face interactions to maximize comprehension and trust.

The findings indicate that participants’ knowledge about their skin condition is significantly influenced by age, gender, prior confusion, and the complexity of their condition. Males showed slightly higher knowledge than females. The studies [9] and [12] report the opposite, with females demonstrating significantly higher dermatology knowledge than males.

Moreover, the results show that participants aged 40–50 years had the highest knowledge levels, whereas those under 20 had the lowest. Importantly, participants who rarely or never experienced confusion about their condition demonstrated substantially higher knowledge compared with those who were often confused. The study [13] highlighted that psychological factors, including confusion, anxiety, or psychiatric comorbidities, can negatively affect patient comprehension. Additionally, individuals with non-complex skin conditions have markedly higher knowledge than those with complex conditions. While education level appeared related to knowledge, this association was not statistically significant. Overall, these results highlight that patient understanding is shaped more by experiential and condition-related factors than formal education. Finally, while the study addresses key factors to improve dermatology patient education, additional studies are required to develop, implement and evaluate approaches that improve patient understanding.

## Conclusion

The present study emphasizes the importance of patient education in dermatology, specifically in the context of the Kingdom of Saudi Arabia, and the significant gaps in patients’ understanding of their skin conditions. There was a considerable proportion of patients that reported some confusion, and less than half of the participants indicated elevated levels of understanding with respect to their skin conditions. The primary factors associated with confusion were limited time with health-care providers and hesitation to ask questions, while patients reported the lowest degree of understanding of their treatment options, the diagnosis, and associated side-effects of treatment. The important findings from the study highlight the need for systematic and targeted educational programs to better inform patients so that they will be better able to understand their skin conditions while also enhancing patient satisfaction and involvement in self-management. It was also found that patients preferred face-to-face consultations to transmit information about their skin conditions, consequently, patients value this personal interaction even in a complicated world with lots of online information. The level of knowledge was even influenced by factors such as age, gender, prior confusion, and complexity, suggesting that direct and experiential learning was more influential to patients understanding their skin condition management versus formal education. Overall, the study articulates that achieving high-quality patient education involves patient education models based on communication strategies as well approaches that reflect individual needs in their management of their skin conditions. This, in turn, will support patient’s achieving better health outcomes and at the same time have a deeper involvement in engagement in their own condition management.

## Data Availability

All relevant data are within the manuscript and its Supporting Information files.

https://docs.google.com/document/d/1xI7yJ_yeVNVQDOeQErPcCS7J6HjWT6iTPt7oAPq4IT0/edit?usp=sharing

## Acknowledgments

We express gratitude to the efforts of data collectors and study participants for their contribution and devotion of time during the data collection period.

The authors have reviewed and edited the output and take full responsibility for the content of this publication.

